# Osteosarcoma: novel prognostic biomarkers using circulating and cell-free tumour DNA

**DOI:** 10.1101/2021.12.12.21267579

**Authors:** Iben Lyskjær, Neesha Kara, Solange De Noon, Christopher Davies, Ana Maia Rocha, Anna-Christina Strobl, Inga Usher, Craig Gerrand, Sandra J Strauss, Daniel Schrimpf, Andreas von Deimling, Stephan Beck, Adrienne M Flanagan

## Abstract

Osteosarcoma (OS) is the most common primary bone tumour in children and adolescents. Despite treatment with curative-intent, many patients die of this disease. Biomarkers for assessment of disease burden and prognoses for osteosarcoma are not available. Circulating-free (cfDNA) and -tumour DNA (ctDNA) are promising biomarkers for disease surveillance in several major cancer types, however only two such studies are reported for OS. In this combined discovery and validation study, we identified four novel methylation-based biomarkers in 171 OS tumours (test set) and comprehensively validated our findings *in silico* in two independent osteosarcoma sample datasets (n= 162, n=107) and experimentally using digital droplet PCR (ddPCR, n=20 OS tumours). Custom ddPCR assays for these biomarkers were able to detect ctDNA in 40% of pre-operative plasma samples (n=72). ctDNA was detected in 5/17 (29%) post-operative plasma samples from patients who experienced a subsequent relapse post-operatively. Both cfDNA levels and ctDNA detection independently correlated with overall survival, p=0.0015, p=0.0096, respectively. Combining both assays increased the prognostic value of the data. Our findings illustrate the utility of mutation-independent methylation-based markers, broadly applicable ctDNA assays for tumour surveillance and prognostication. This study lays the foundation for multi-institutional collaborative studies to explore the utility of plasma-derived biomarkers for predicting clinical outcome of OS.

## 1. Introduction

Osteosarcoma (OS) is the most common primary malignant bone tumour in childhood and adolescents, generally arising in the metaphysis of long bones^1^. Approximately 80-90% of all OS tumours are high grade, and despite toxic multimodal therapies, survival for the majority of patients with OS has not improved over the last 40 years^2^, with 30-40% of patients with OS dying of their disease^3^. Although, response to chemotherapy is an indicator of survival^4^, it is currently not possible to predict which patients are likely to respond to chemotherapy.

Studies have demonstrated the value of minimally invasive surveillance assays in cancer management including assessment of the efficacy of treatment through detection of residual tumour following surgery, and predicting response to chemotherapy or disease relapse^5^. Measurement of total circulating free and tumour DNA (cfDNA and ctDNA, respectively), representing DNA shed into the bloodstream by tumour cells, has been shown to correlate with important clinical endpoints (*e*.*g*. progression-free survival and or overall survival) in several cancer types, as reviewed in^6–8^. Barris *et al*. published the first ctDNA study in OS patients (10 patients, 28 samples) using targeted sequencing of genetic alterations detected in the tumours^9^, and found that ctDNA was detected in approximately half of the plasma samples (46%)^9^. Shulman *et al*. used ultra-low-pass whole-genome sequencing (WGS) and detected ctDNA in 57% (41/72 OS patients) of localised OS prior to treatment, and reported that detection of ctDNA was associated with inferior survival but the results were not statistically significant^10^.

In the human genome, DNA methylation predominantly occurs at CpG dinucleotides, the site where a cytosine residue precedes a guanine residue in the 5’-3’ direction. In the healthy human genome the majority of CpGs are methylated except at CpG islands^11^. Conversely, a cancer genome is often characterised by global hypomethylation and focal hypermethylation at CpG islands, leading to both genomic instability and transcriptional repression^12^. This feature of cancer has been exploited by studies, which demonstrate ctDNA methylation profiles as a highly sensitive and specific means of detecting cancers^13^. To date, only a few studies have investigated methylation of OS^14–16^. The use of DNA methylation as a marker of ctDNA in OS is attractive because this tumour is characterised by complex genomes and high inter-tumour heterogeneity rather than recurrent genetic alterations^17–19^, making selection of a universal genetic ctDNA marker challenging. Furthermore, as changes in methylation represent an early event in cancer and are considered to be retained through tumour evolution^20^, they could be exploited as powerful biomarkers and would be expected to be only moderately confounded by tumour genetic heterogeneity^21^. For this reason, this approach potentially has advantages over the two previous ctDNA studies in patients with osteosarcoma in which genomic alterations were targeted^9,10^.

It has been previously established that high levels of DNA in the circulation (cfDNA) have been linked to inferior outcome^22–24^. However, in contrast to ctDNA which represents a tumour-specific assay, cfDNA levels can be influenced by other factors including physical activity, age and rate of clearance. For these reasons cfDNA is considered a less reliable biomarker, particularly when measured shortly after surgery^6^. Despite this, the clinical utility of cfDNA as a prognostic marker has been proven in a wide range of cancers^6^.

The aim of this study was to identify plasma-derived biomarkers in patients with OS that could be used for predicting outcomes.

## 2. Materials and methods

### 2.1. Ethical Approval

The Royal National Orthopaedic Hospital (RNOH) Biobank is approved by the National Research Ethics Committee of the Health Research Committee (reference: Integrated Research Application System (IRAS) project identifier: 272816). This study was approved by the National Research Ethics Committee approved UCL/UCLH Biobank Ethics Committee (project no: EC17.14).

### 2.2. Training methylation cohort

Our *in-house* dataset consisted of 750 samples of various sarcoma subtypes (Supplementary Table 1) (EMBL-EBI, under accession number E-MTAB-9875) previously published^25^. Infinium 450K methylation data was downloaded from TCGA (https://www.cancer.gov/tcg) using TCGAbiolinks^26^ and from Marmal-aid^27^ (kindly provided by Dr Robert Lowe, UCL, QMUL, UK) (Supplementary Table 2).

### 2.3. External validation cohorts for identified methylation markers

Validation Set I consisted of 450K and EPIC methylation raw IDAT files from 162 OS from Heidelberg, Germany (provided by DS and AVD^28^, GSE140686) and 699 peripheral blood leucocytes (PBLs) from non-cancer patients (GSE125105^29^). Validation Set II consisted of 21 publicly available OS samples (GSE58770^30^), 86 OS samples from the TARGET-OS (Children’s Oncology Group and The Hospital for Sick Children in Toronto, Canada, dbGaP accession phs000218.v21.p7, http://ocg.cancer.gov/programs/target and 732 healthy blood samples (GSE87571^31^).

### 2.4. Biomarker discovery and digital droplet PCR assay design

To avoid false positive results in the plasma samples, we first excluded all CpG sites that showed signs of DNA methylation (β-value > 0.2) in more than 1% of all peripheral blood leucocytes (PBLs) samples (TCGA/Marmal-aid cohorts). Next, we excluded a) CpGs that were hypomethylated (β < 0.5) in more than 20% of all OS cases, b) hypermethylated CpGs (β > 0.5) that were detected in more than 50% of other cancers and c) hypermethylated CpGs (β > 0.2) detected in more than >30% of healthy tissues (Supplementary Figure 1B).

The 17 identified candidate methylation markers (CpGs) were annotated using the *COHCAP*^32^ R package (using the hg19 genome build). Methylation-specific digital droplet PCR (ddPCR) assays were designed using Primer3Plus^33^ and MethPrimer^34^, and checked using BiSearch^35^. Receiver operating characteristics (ROC) analyses were performed using the pROC R package^36^ to determine the sensitivity and specificity of the probes both individually and in combination (where at least two of the markers were positive). A β-value of 0.5 was chosen as the threshold for determining if a sample should be classified as positive. To be able to use a droplet digital PCR assay with the available plasma (1-2 mL), we reduced the number of markers from 17 to four. This was achieved by first calculating the sensitivities and specificities in the Training and Validation sets (Supplementary Table 8) and then identifying regions for which ddPCR assays could be designed. This led us to selecting the following CpGs for experimental validation; cg02169391, cg22082800, cg25680486, and cg26100986.

### 2.5. Clinical samples for experimental validation

Three clinical sample sets were available for experimental validation: 1) high grade OS tumour samples from 20 patients (Supplementary Table 3), 2) control plasma samples obtained from 47 healthy volunteers (2.5 mL plasma) and 69 individuals with benign bone disease (see below for list) and degenerative joint disease (referred to hereafter as non-cancer patients; 4.5 mL plasma, Supplementary Table 4), and 3) samples from 72 patients with OS; these included blood for germline DNA and their pre-operative, pre-treatment plasma samples, in addition to 17 post-OP plasma samples (a total of 89 plasma samples with a median of 4.5 mL, range 3-4.5 mL (1-2 mL were used for ddPCR analysis), overview in Supplementary Table 6). The samples of benign bone disease mentioned above included simple bone cysts, osteoarthritis, osteoblastoma, fibrous dysplasia, osteoid osteoma, osteochondroma, tenosynovial giant cell tumour, synovial chondromatosis; the degenerative joint disease represented patients undergoing hip or knee replacements (Supplementary Table 7).

#### Clinical correlates

Osteosarcoma patients were identified from a research database of patients diagnosed and managed within the London Sarcoma Service and included patients of all ages with demographic data incorporating primary site of disease, tumour grade and size, treatment parameters (chemotherapy, surgery and radiotherapy) as well as pathological response to chemotherapy and outcome (Supplementary Table 5).

### 2.6. Statistical analysis

All statistical analysis was performed in R (version 3.6.1)^37^. A p-value <5% was deemed statistically significant. A ctDNA-positive sample was defined as a sample with a least two of the four methylation markers above the limit of detection (LOD). Survival analysis was performed using the Kaplan-Meier estimator with death as the end point. The multivariate analyssiis wass a Cox proportional hazards regression model with time from diagnosis to death or censorship (time to last follow-up) as an end point.

### 2.7. Data availability

*In-house* raw methylation array data have previously been published^25^ and are available from the ArrayExpress database at EMBL-EBI (www.ebi.ac.uk/arrayexpress), accession number E-MTAB-9875.

## 3. Results

### 3.1. *In silico* identification and validation of methylation markers through large scale bioinformatic analyses

Our first step in the identification of novel CpG sites as candidates for ctDNA assays involved downloading and interrogating the publicly available methylation data from 8,730 and 2,885 human tissue samples from the TCGA^38^ and Marmal-Aid^27^, respectively (Supplementary Table 2, and Figure 1 for study outline). 10,766 samples were eligible for further analysis after filtering for incomplete data (Methods, Supplementary Figure 1A), and were analysed alongside 750 in-house sarcoma methylation profiles (Supplementary Figure 1A). Data from the remaining 10,766 samples were used for bioinformatic analysis together with 750 of our in-house sarcoma samples^25^. Hence, the final discovery group included methylome data from 11,516 samples, including 1,578 blood samples, 202 normal tissues, 9,565 non-OS cancer samples and 171 OS tumour samples (Supplementary Figure 1, Supplementary Tables 1 and 2).

**Figure 1:**
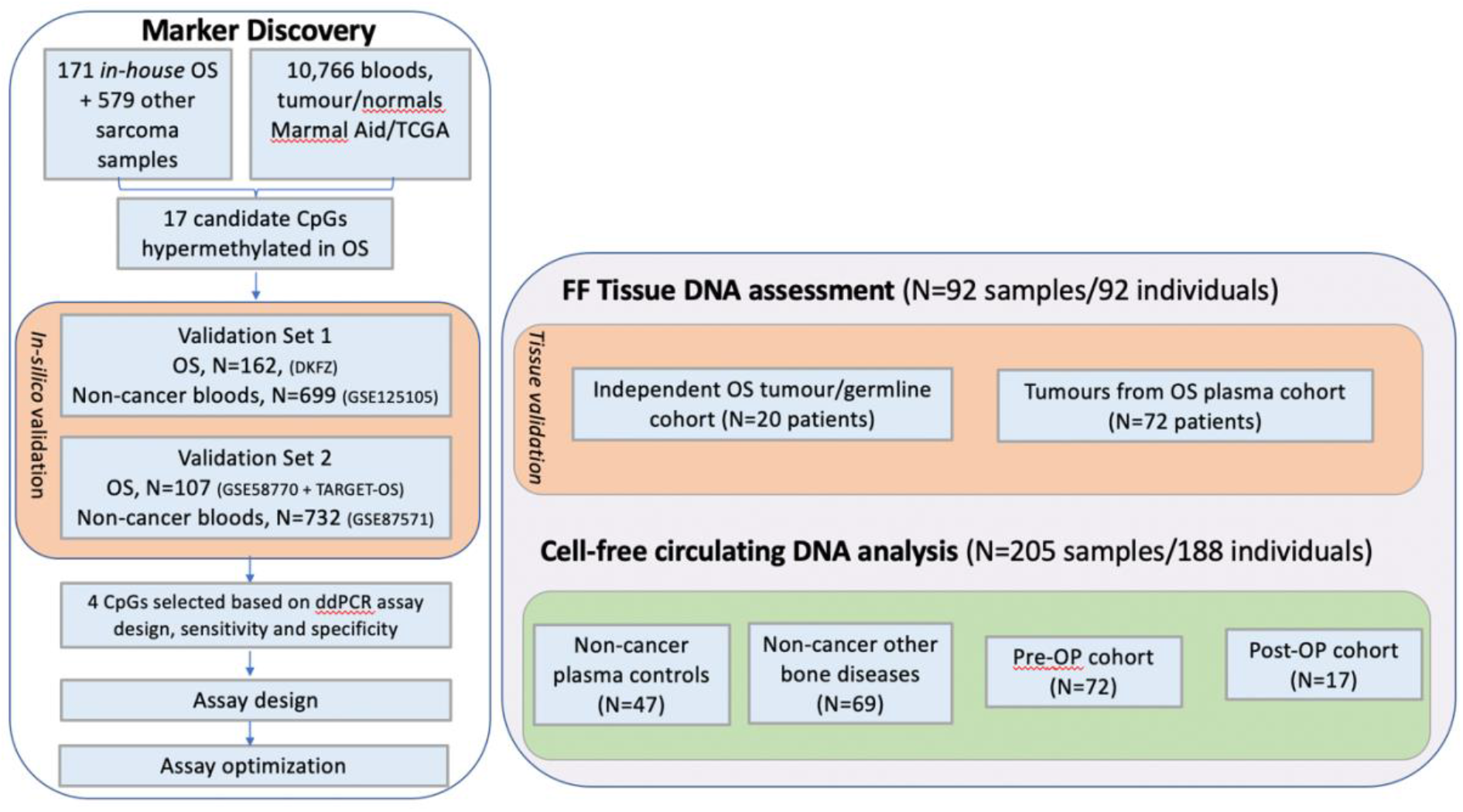
Overview of the combined biomarker and validation study. 17 osteosarcoma-specific candidate methylation markers were identified through a stepwise approach for selecting CpG sites uniquely methylated in OS (see Methods). Four markers were chosen for methylation-specific digital droplet PCR (ddPCR) analysis. The markers were subsequently validated both *in silico* and experimentally in tissue, before being used for circulating tumour DNA assessment of plasma samples from osteosarcoma patients. ‘Other sarcoma’ denotes various other non-OS soft tissue and bone sarcoma subtypes. FF: fresh frozen tumour tissue.

#### 3.1.1. Osteosarcoma-specific DNA methylation markers

We next took a stepwise approach to identify DNA methylation markers (CpG sites), which were uniquely methylated in OS (Supplementary Figure 1B). Using our screening pipeline (Supplementary Figure 1B), we identified 17 candidate hypermethylated markers for OS (Supplementary Figure 2). To obtain an estimate of their combined sensitivity and specificity, we performed receiver operating characteristic (ROC) curve analysis on 171 OS and 1,578 non-cancer PBLs (Training Set). The mean area under the curve (AUC) was 0.994 and included all 171 OS and only 11/1578 (0.7%) non-cancer PBLs samples (Supplementary Figure 3A).

#### 3.1.2. Candidate DNA methylation markers show robust performance in independent OS datasets

Two independent rounds of *in silico* validation of these candidate methylation markers were performed using Validation Set 1 comprising 162 OS tumour and 699 non-cancer PBLs, and Validation Set II consisting of 107 OS tumour and 732 PBL samples (Materials and Methods). By including all 17 candidate markers with a β-value cut-off ≥ 0.5, all OS in both validation sample sets were correctly classified. None of the 699 PBLs were hypermethylated (β ≥ 0.5) at any of the 17 candidate markers in Validation Set 1, and only two of 732 PBLs (0.27%) from non-cancer patients were misclassified as OS in Validation Set 2. ROC analyses gave an AUC = 1.00 for Validation Set 1 and AUC = 0.99 for Validation Set 2 (Supplementary Figure 3B and 3C).

#### 3.1.3. Four candidate DNA methylation markers were selected for experimental validation

Digital droplet PCR-based assays were chosen as the method for ctDNA measurement as this represents a widely employed and highly sensitive, specific, and cost-effective methodology. The amount of plasma available from patients and the number of markers that can be multiplexed represent limitations of this approach and therefore from the 17 candidate markers, we selected four CpGs (cg02169391, cg22082800, cg25680486, and cg26100986) to take forward (Methods). Hypermethylation of all four markers correctly identified 421/440 of all OS samples studied (95.7%) and misclassified only 6/3009 (false positives = 0.2%) non-cancer PBLs as OS *in silico* (Training and the two Validation Sets) (Supplementary Table 8, Supplementary Figure 4).

### 3.2. Experimental validation of methylation markers in clinical OS cohorts

#### 3.2.1. Candidate markers are highly sensitive and specific

To validate further the sensitivity and specificity of the four markers, we tested these as two duplex ddPCR assays experimentally on samples from another set of 20 high-grade OS and matching germline blood DNA (Supplementary Table 3). Hypermethylation at these CpG sites was confirmed in each of the tumours and in none of the germline samples (Supplementary Figure 5).

To refine the limit of detection (LOD, Materials and Methods and Supplementary Methods), we also applied the four assays (two duplexed assays) to 47 control plasma samples from patients with non-neoplastic conditions (Supplementary Table 6, Supplementary Figure 5) and plasma from 69 individuals with benign bone disease (Methods, Supplementary Table 7). 3/116 samples gave a false positive result (cases 57920 and 59831 (osteochondromas) and 63637 (healthy volunteer), with an overall specificity of 97.4%.

#### 3.2.2. Circulating tumour DNA detection correlates with disease progression

Finally, we measured the four methylation markers in matched tumour, PBLs and pre-treatment plasma samples from 72 patients with OS (Supplementary Table 4 and 5). Based on the probes with the highest methylation levels observed in the tumour tissue, we ran one duplexed assay on the matched plasma sample. If neither or only one of the methylation markers was detected, we ran the other duplex assay. Although we observed methylation of at least one marker in 50/72 (69%) samples, we set a minimum of at least two markers to be detected above the LOD for a sample to be classified as ‘ctDNA positive’ and employed this criterion hereafter.

Clinical outcome data was then assessed against our pre-operative ctDNA assay results (Table 1). Multivariate analysis confirmed that the presence of metastases at diagnosis is an independent prognostic factor in our cohort (Table 2), a finding consistent with the literature^39^. ctDNA assays were positive in 29/72 (40%) pre-operative samples (ctDNA^pre+,^ Table 1, Supplementary table 4). Within this ctDNA^pre+^ group, 12/29 patients had metastatic disease at diagnosis. Of the 17 ctDNA^pre+^ patients with localised disease at diagnosis, 10 suffered a disease related event (death, n= 8; or metastasis, n=2). The remaining seven ctDNA^pre+^ patients were alive without disease at last follow up (median follow-up = 5.3 years, range 0.9-11.3 years, Supplementary Table 5). Overall, the detection of pre-treatment ctDNA correlated with inferior survival outcomes (p<0.01, Figure 2A, Table1) and with a disease-related event (p=0.01, chi-square test), but not with tumour size (p=0.14, Student’s t-test), gender (0.53) or age at presentation (p=0.92). ctDNA positivity did not have a significant association with outcome on multivariate analysis, however it should be noted that this model also included metastatic disease status at diagnosis which is known to be linked to increased tumour burden^40,41^, and thus higher probability of ctDNA detection.

**Table 1.** Correlation of clinical outcome with pre-treatment ctDNA. PR: poor responder; GR: good responder, NCG: No chemo given, ctDNA: circulating tumour DNA.

|  | ctDNA pre<br>neg (n=43) | ctDNA pre pos<br>(n=29) |
| --- | --- | --- |
| Metastasis at<br>diagnosis<br>(n=19) | 7 (16%) | 12 (41%) |
| No<br>metastases at<br>diagnosis<br>(n=53) | 36 (68%) | 17 (32%) |
| Die of disease<br>(n=32) | 14 (33%) | 18 (62%) |
| Disease-related event<br>(n=39) | 18 (42%) | 21 (72%) |
| Response to<br>chemotherapy | PR: 28<br>(65%)<br>GR: 9<br>(21%)<br>NCG:6<br>(14%) | PR: 11 (38%)<br>GR: 8 (28%)<br>NCG:4 (14%)<br>Unknown/NA: 5 |

**Table 2:** Uni-and multivariate analysis of clinical factors and their relation to survival. cfDNA: circulating free DNA; ctDNA: circulating tumour DNA; pre-OP: pre-operatively.

| SURVIVAL | Univariate analysis |  |  | Multivariate analysis |  |  |
| --- | --- | --- | --- | --- | --- | --- |
|  | HR | 95% CI | P-value | HR | 95% CI | P-value |
| **Pre-OP cfDNA (n=72)** |  |  |  |  |  |  |
| <75th centile (n=53) | 0.34 | 0.38 | **0.005** | 0.56 | 0.43 | 0.18 |
| >75th centile (n=19) | REFERENCE |  |  |  |  |  |
| **Pre-OP ctDNA (n=72)** |  |  |  |  |  |  |
| Neg (n=43) | REFERENCE |  |  |  |  |  |
| Pos (n=29) | 2.51 | 0.36 | **0.01** | 1.48 | 0.42 | 0.36 |
| **Metastasis at**<br>**presentation (n=72)** |  |  |  |  |  |  |
| Yes (n=19) | 5 | 0.37 | **1.2\*10<sup>-5</sup>** | 3.78 | 0.40 | **0.0096** |
| No (n=53) | REFERENCE |  |  |  |  |  |
| **Gender (n=72)** |  |  |  |  |  |  |
| Male (n=38) | REFERENCE |  |  |  |  |  |
| Female (n=34) | 1.16 | 0.36 | 0.68 |  |  |  |
| **Age (n=72)** |  |  |  |  |  |  |
| <30 years (n=62) | REFERENCE |  |  |  |  |  |
| =>30 years (n=10) | 1.14 | 0.54 | 0.81 |  |  |  |
| **Max tumour size (n=64)** |  |  |  |  |  |  |
| <=80 mm (n=14) | REFERENCE |  |  |  |  |  |
| >80 mm (n=50) | 1.38 | 0.5 | 0.51 |  |  |  |

**Figure 2:**
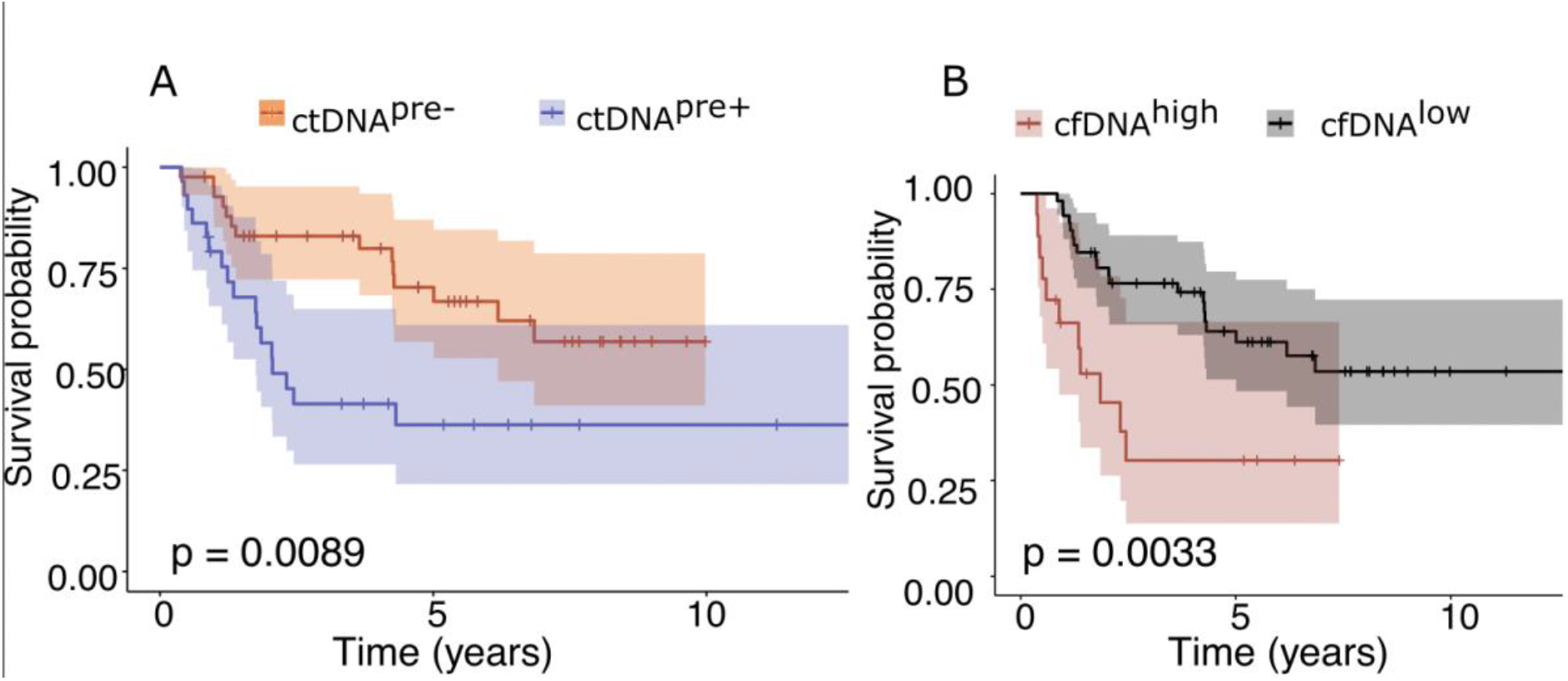
Detection of ctDNA or high levels of cfDNA pre-operatively correlates with survival. A) Detection of ctDNA and **B)** cfDNA pre-operatively correlate with overall survival (disease-related mortality or censorship). Survival analysis utilised a standard Cox proportional hazard model. The plots are censored after 12 years.

We also analysed post-operative plasma available from 17 of the 72 patients with OS in whom ctDNA was detected pre-treatment. ctDNA was detected in 5/17 (29%, median days from surgery 18, range: 7-30 days), 4/5 of whom developed recurrent disease (local recurrence or metastasis) and or died of disease (Supplementary Table 5), the remaining patient (OS_66) is alive and well nine years post-surgery. The ctDNA biomarkers were detected in 2/4 patients before the clinical relapse was identified. Detection of ctDNA post-operatively was not associated with the presence of metastasis at diagnosis (p=0.45, Student’s t-test). Of the patients for whom post-operative samples were available, four and 12 resection specimens were reported as having a good and poor response to chemotherapy, respectively (one unknown), and all cases were reported as having been fully excised.

#### 3.2.3. Total circulating free DNA levels correlate with outcome

Finally, we measured the total amount of cfDNA in the patients’ plasma samples, the median level of which was 1,848 copies/mL plasma (range 231-15,180 copies/mL plasma). This correlated with survival (cutoff=75^th^ centile, cfDNA^high^, p=0.003, Figure 2B), but not with tumour size (rho=0.63, spearman rank test), metastasis at diagnosis (p=0.18, Kruskal-Wallis rank sum test) or anatomical site (femur versus tibia; p=0.63, Kruskal-Wallis rank sum test). 12/19 (63%) of those patients classified as cfDNA^high^ were also ctDNA^pre+^; of the remaining 7/19 cfDNA^high^ cases without ctDNA^pre+^, two had metastasis at diagnosis and another two relapsed (Supplementary Table 5). Including cfDNA^high^ as a risk factor for adverse outcomes alongside ctDNA^pre+^ and metastases at diagnosis resulted in 41/72 (57%) patients classed as high risk at the time of diagnosis. This three risk factor approach correlated with a poor prognosis for this group (p=0.01, Supplementary Figure 6, Supplementary Table 5).

## 4. Discussion

The standard of care for patients with osteosarcoma^2^ results in an approximate 70% five-year overall survival, thus despite aggressive multi-drug treatment 30-40% die of their disease. The identification of metastases at diagnosis is the most reliable indication that a patient with osteosarcoma has an unfavourable prognosis. However, other factors also contribute to outcome, including tumour size and site, and response to chemotherapy, site^42,43^. Nevertheless, even when combining all these clinical variables, clinicians lack sufficient information to stratify patients adequately for treatment and to offer patients with OS a reliable prognosis.

Here, we report a pipeline for the development and application of DNA methylation markers in OS and demonstrate their utility in detecting ctDNA and show that their detection in plasma correlates with inferior clinical outcome. Importantly, these methylation markers were detected in the pre-treatment ctDNA of 32% of patients who did not have metastases at presentation, indicating that this assay has the potential to complement and add value to current criteria for guiding treatment and providing more accurate prognoses to patients. However, 24% of patients with a positive pre-operative ctDNA test remain disease-free, which may in some cases be linked to a follow-up period of less than five years. The cfDNA results confirmed the ctDNA findings in most cases with 63% of patients with a high level of cfDNA also being ctDNA^pre+^.

The four selected methylation markers appear to be highly specific and sensitive for OS, being detected in 421/440 (96%, Discovery and Validated Sets) cases analysed *in silico*, and therefore have the potential to be an applicable test for up to 90% of patients with high grade OS. False positive ctDNA results in patients without OS using our methylation markers were rare (2.6%) and should not lead to misdiagnoses or inappropriate management provided results are interpreted in the context of the full clinical picture including medical imaging typically in the form of a multidisciplinary meeting. In this context, our study represents a first step towards the goal of turning cancer including OS into a chronic but manageable disease^44^.

The reason for failing to detect ctDNA in 60% of patients, particularly in those presenting with metastases or whose disease progressed, is unexplained. Nevertheless, it is noteworthy that relatively similar results (54%^9^ and 43%^10^ not detected) were reported in two other studies, which involved different study designs and which employed different technologies for the detection of ctDNA. The study of Barras *et al*. only included ten patients, and that of Shulman *et al*. only studied patients without metastasis at presentation^9,10^. Furthermore, our study included patients aged between five and 95 years, whereas the other two studies were conducted in patients younger than 35 years^9,10^. It is possible that the sensitivity of all assays undertaken in the different studies could be improved if larger blood volumes were available for analysis. However, the similar findings across the three studies suggest that the pathobiology of tumours that shed detectable levels of ctDNA may be different from those where this does not occur. This finding is not comparable to the results from other cancers, where ctDNA is detectable in the vast majority of high grade cases in cancers such as lung^45^, ovary ^46^ and bladder^47^. Indeed, ctDNA is even detected in small colorectal cancer lesions and is being considered as a screening test for early detection of this disease^48^. ctDNA is also detected in virtually all high grade chondrosarcomas, another bone cancer, but not in low grade cases^49^.

It would be interesting to test head-to-head the various technologies which have been employed to assess ctDNA in OS patients, but this is difficult to undertake because each assay requires a considerable volume of blood. Nevertheless, a greater understanding of why ctDNA is only detected in some patients may be revealed by interpreting the findings in the context of other features such as the pathology, the medical imaging, as well as multi-omic tumour profiles.

The limitations of the study are related to the application of the methylation markers to clinical outcome and are largely attributable to the rarity of osteosarcoma, the incidence of which is 0.27/100,000 per year in England^50^. The consequence of this includes the relatively small number of samples analysed within a heterogenous cohort of patients across all age groups, and a relatively short median follow-up of approximately 3.5 years. The small number of post-operative samples, and the absence of an independent cohort in which our findings could be validated are also limitations. The small numbers of samples that were available for analysis post-treatment was disappointing but the finding that ctDNA is detectable prior to presentation of clinical relapse is encouraging and requires further investigation.

The study highlights the need for multi-centre collaboration, to enable the prospective systematic collection and sharing of patient samples with annotated clinical data, to recruit sufficient patient numbers for validation of these biomarkers and determination of their clinical utility in osteosarcomas, or any other rare disease. Multi-centre studies are fraught with challenges; they are expensive, time-consuming to establish, and it can be difficult to agree and prioritise research aims and studies. Nevertheless, results generated by different groups can add value and provide an opportunity for cross-validation. To this end, the ICONIC study (Improving Outcomes through collaboration in Osteosarcoma)^51^, brought together the osteosarcoma clinical and research communities across the UK and provides the infrastructure to prospectively evaluate clinical and biological questions, such as this, to better stratify patients for treatments and improve outcome. It is currently recruiting newly diagnosed OS patients across the UK^52^.

## Data Availability

l data produced in the present study are available upon reasonable request to the authors.

https://www.ebi.ac.uk/arrayexpress/experiments/E-MTAB-9875/

## Acknowledgements

Funding for this project was received from the Tom Prince Cancer Trust, The Rosetrees and Stoneygate Trusts (M46-F1) and the Bone Cancer Research Trust (BRCT: 6519). IL is supportedby the Lundbeck Foundation (R303-2018-3018) and previously by the Tom Prince Cancer Trust. NK was funded by Imperial College London. SB acknowledges support from the Wellcome Trust (WT-218274/Z/19/Z). The study was also supported by the National Institute for Health Research, UCL Genomics and Genome Engineering, UCL Great Ormond Street Institute of Child Health and Pathology Core facilities, the Cancer Research UK University College London Experimental Cancer Medicine Centre, and the RNOH Research and Development Department. AMR and CD was supported by the Tom Prince Cancer Trust. SJS was supported by the National Institute for Health Research, the University College London Hospitals Biomedical Research Centre.

The authors thank Dr Sarah Ø. Jensen and Dr Jacob H. Fredsøe, Aarhus University in Denmark, for inputs to the biomarker discovery, Professor Gert Attard and Dr Anjui Wu for a valuable discussion of the experimental design as well as Dr Robert Lowe for sharing data from the Marmal-Aid database. The results generated in this article are partly based upon data generated by the TCGA research Network (http://cancergenome.nih.gov), Marmal-Aid database and the Therapeutically Applicable Research to Generate Effective Treatments (TARGET) initiative managed by the NCI. We acknowledge the RNOH Biobank Team and all healthcare workers who cared for the patients. We give a special thanks to the patients for their donating invaluable samples for research.

## Statement of author contribution

Conceptualization: IL, SB, AMF; Sample curation: AMF; Data curation: IL, NK, AMR, CD, ACS, IU, DS, AVD; Analysis: IL, NK, SDN; Clinical data: AMF, SDN, CG, SJS; Supervision: SB, AMF; Writing: IL, SB, AMF, SDN; Review and editing: All authors.

## Notes

### Competing Interest Statement

The authors have declared no competing interest.

