## Supplementary Figure for "Osteosarcoma: novel prognostic biomarkers using circulating and cell-free tumour DNA"

**<u>Supplementary Figures</u>**

Supplementary Figure 1. Overview of included samples (TCGA, marmal-aid, in-house)
Supplementary Figure 2. Plot for each of the 17 candidate CpGs
Supplementary Figure 3. Discovery cohort and validation cohorts for the 17 potential markers
Supplementary Figure 4. The designed assays detect minimal methylation levels in non-cancer plasma cfDNA samples
Supplementary Figure 5. Specificity assessed in a non-cancer cohort (n= 69) consisting of related bone disease.
Supplementary Figure 6. Combining metastasis at diagnosis, ctDNApre+ and cfDNAhigh predicts survival (‘pos_marker’).

**Supplementary Figure 1. *In silico* analysis to identify potential osteosarcoma-specific methylation markers.** A) Overview of the samples analysed. B) Overview of the exclusion of probes. We excluded CpGs hypermethylated in blood, other cancers or in normal tissue and excluded CpGs hypomethylated in osteosarcoma tumour samples.

**A**

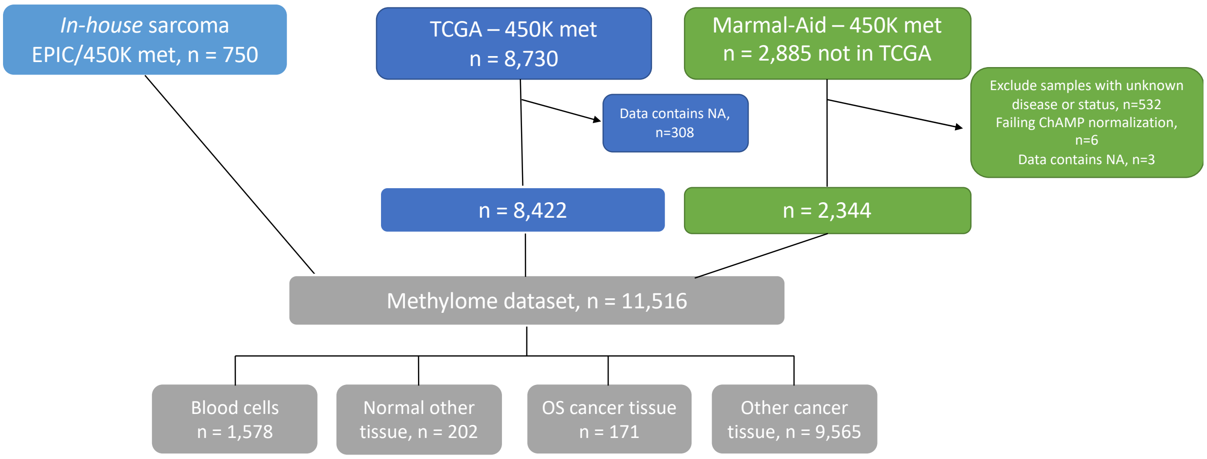


**B**

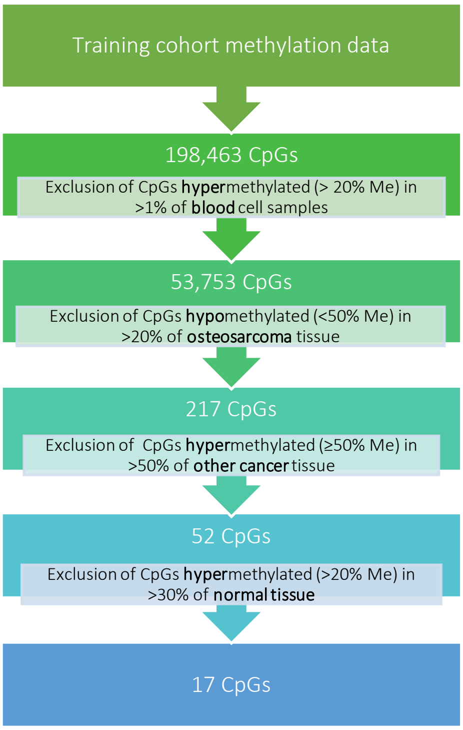

**Supplementary Figure 2. Plot for each of the 17 potential CpGs**. The plots show the beta value (y axis) for each of the 17 potential CpGs for osteosarcoma tumours, blood samples, normal tissue and other cancers (not osteosarcoma, but other sarcomas are included). An * indicate the four CpGs chosen for experimental validation.

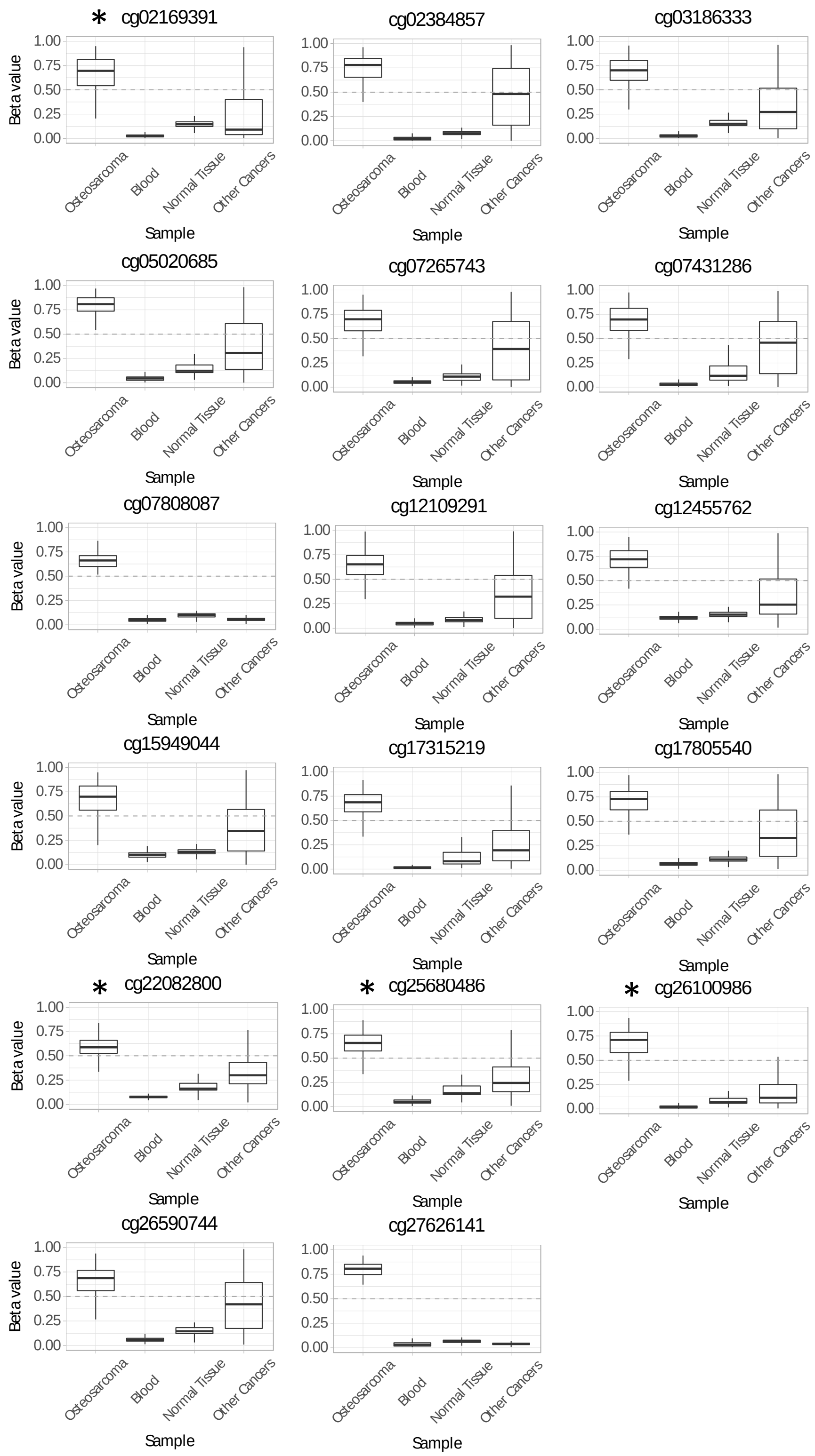

**Supplementary Figure 3**. Discovery cohort and validation cohorts for the 17 potential markers. To get an estimate of the sensitivity and specificity of the 17 candidate methylation markers, we performed receiver operating characteristic (ROC) curve analysis for the A) discovery cohort consisting of 171 OS and 1,578 non-cancer PBLs. The mean area under the curve (AUC) was 0.994; none of the 171 OS samples were misclassified whereas 11/1578 (0.70%) non-cancer PBLs samples were misclassified. B) Validation cohort I: 162 OS tumour samples from our collaborators in Heidelberg and an online-available dataset containing 699 PBLs from non-cancer patients (GSE125105). C) Validation cohort II: 107 OS tumours (GSE58770 and TARGET-OS) and 732 PBLs from non-cancer patients (GSE87571).

**A) Discovery cohort**

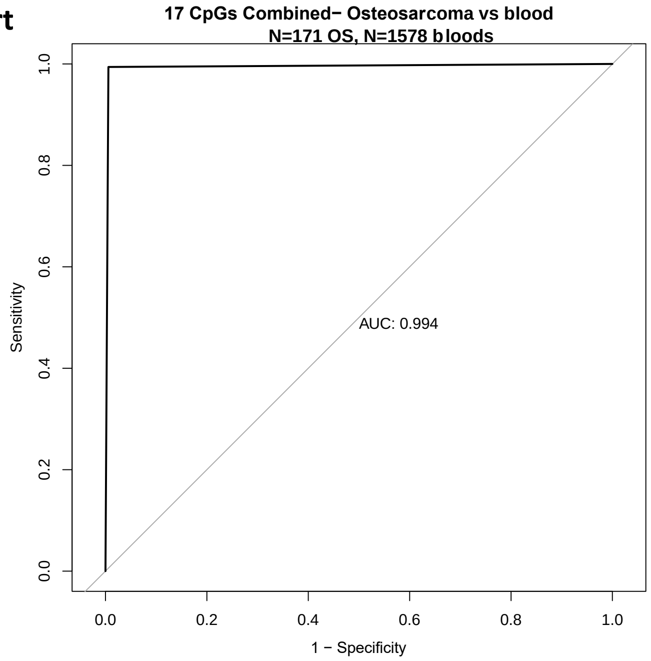


**B)** Validation cohort I

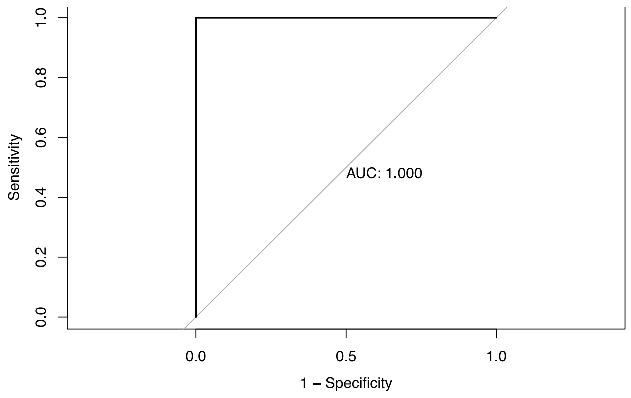


**C)** Validation cohort II

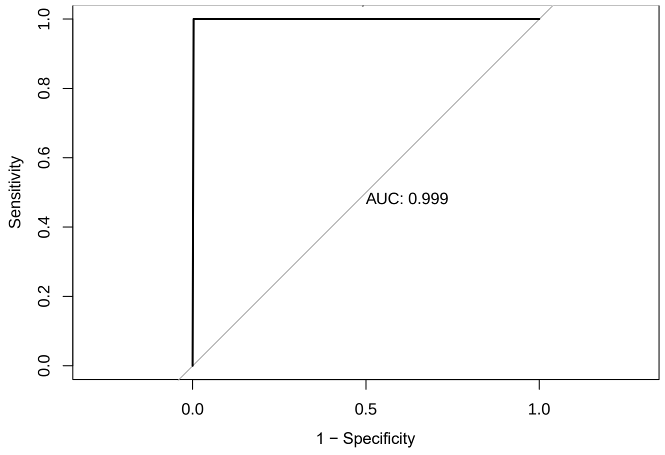

**Supplementary Figure 4.** ROC analyses of the 4 CpG marker panel in the training Set and two independent Validation Sets.
ROC probability curves generated using the training set data (A), and validation Set 1 (B) and 2 (C), with the AUC (area under curve), sensitivity (Sens.) and specificity (Spec.) values displayed on each plot. Plots were generated using the pROC package in R.

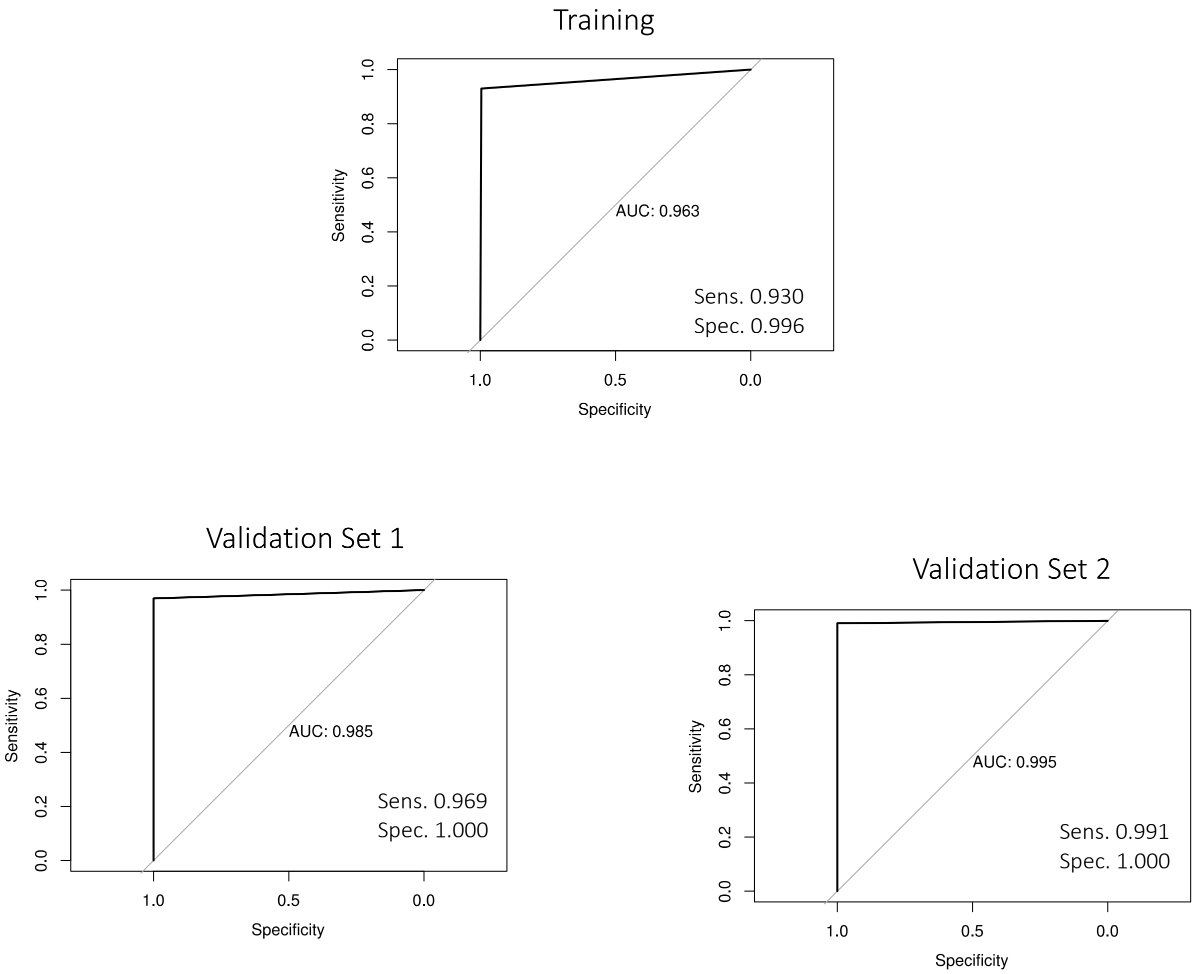

Supplementary Figure 5. **The designed assays detect minimal methylation levels in non-cancer plasma cfDNA samples.**

A display of boxplots with underlaid scatter plots to show CpG methylation levels (cg02169391 (**A**), cg22082800 (**B**), cg25680486 (**C**), cg26100986 (**D**) calculated by each ddPCR CpG assay using non-cancer plasma control samples (n=47) and OS tumour samples (n=20). The numbers displayed on the panel represent the average methylation level of each sample group. The coloured dashed lines represents optimal LOD thresholds (cg02169391 – 10.0%, cg22082800 – 3.0%, cg25680486- 3.0%, cg26100986 – 3.0%).

The Mann-Whitney U Test was used to compare the methylation levels in OS tumour and non-cancer plasma samples for each marker. **** represents p<0.0001.

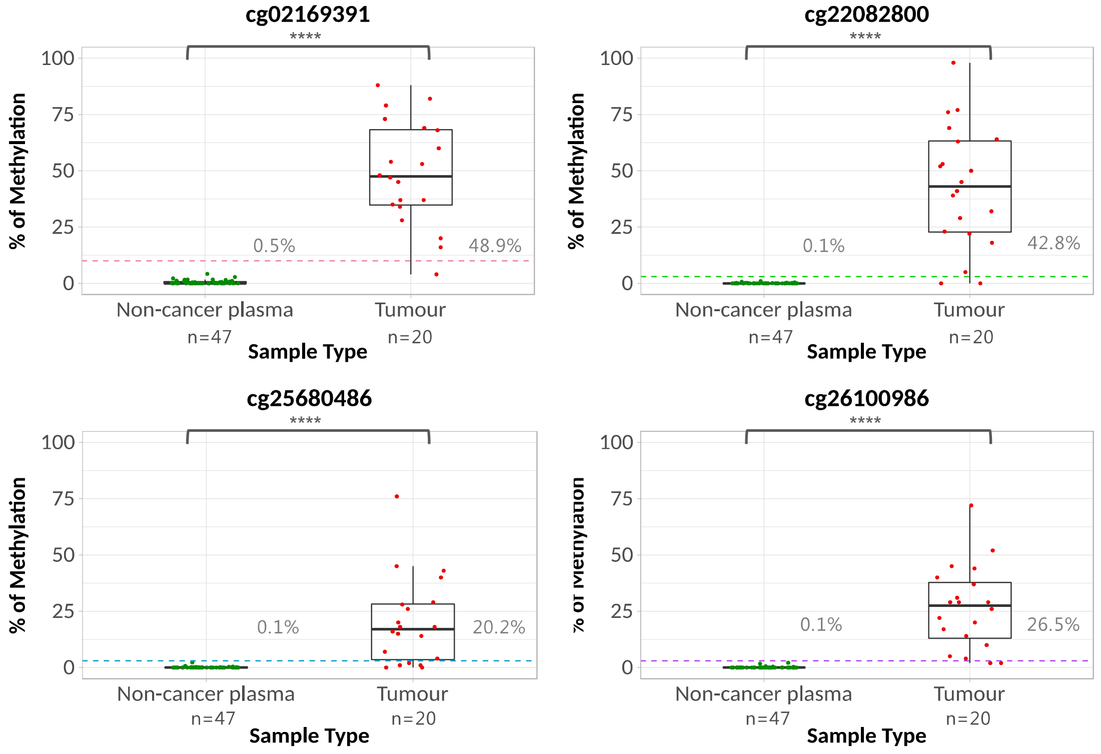

**Supplementary Figure 6.** Combining metastasis at diagnosis, ctDNA$^{pre+}$ and cfDNA$^{high}$ predicts survival (‘pos_marker’). Group ‘neg_marker’ represents those in whom ctDNA was not detected pre-operatively, who did not have metastases at diagnosis nor had a high level of cfDNA.

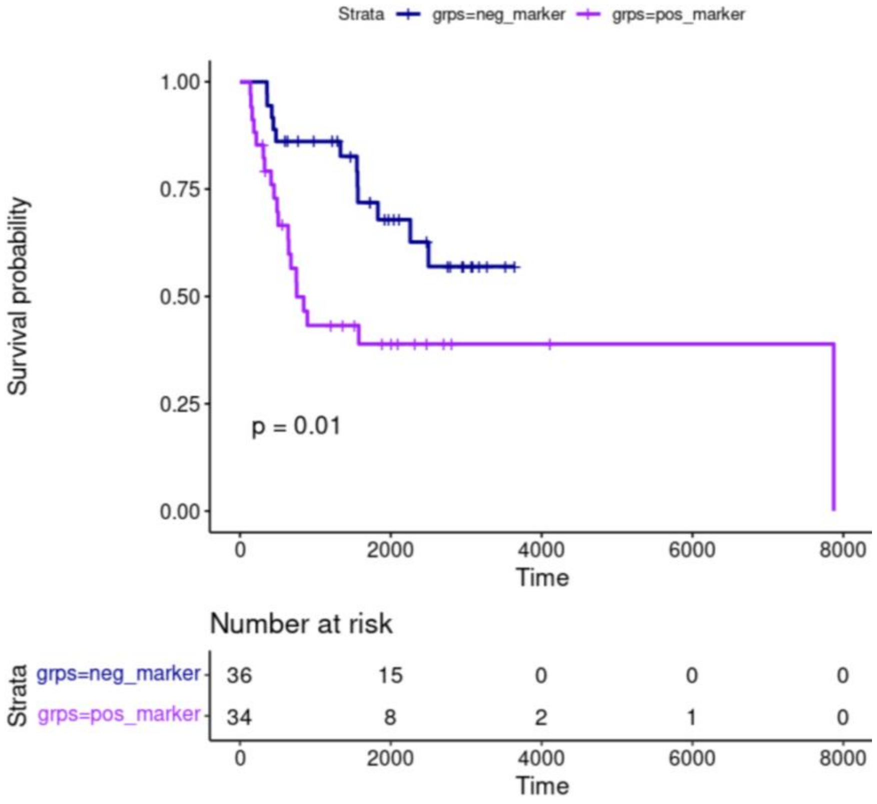
