## Supplementary Table 1 for "Osteosarcoma: novel prognostic biomarkers using circulating and cell-free tumour DNA"

Supplementary Table 1 – *in-house* dataset (450K and EPIC array data)

| Cancer type | Number of cases |
| --- | --- |
| Adamantinoma | 14 |
| Osteosarcoma | 188 |
| Alveolar soft part sarcoma | 11 |
| Angiosarcoma | 9 |
| Atypical neurofibroma | 6 |
| Aneurysmal bone cyst | 12 |
| Angiomatoid fibrous histiocytoma | 1 |
| Carcinoma | 2 |
| Chondroblastoma | 16 |
| Chondromyxoid Fibroma | 14 |
| Chondrosarcoma | 116 |
| Chordoma | 49 |
| Giant cell tumour of bone | 42 |
| Epithelioid sarcoma | 7 |
| Ewing's Sarcoma | 1 |
| Fibrous Dysplasia | 6 |
| Infantile fibrosarcoma | 1 |
| Leiomyosarcoma | 6 |
| Liposarcoma | 16 |
| Malignant peripheral nerve sheath tumour | 90 |
| Myxofibrosarcoma | 5 |
| Myxoinflammatory fibroblastic sarcoma | 3 |
| Myxoma | 1 |
| Non-ossifying Fibroma | 13 |
| Osteoblastoma | 13 |
| Osteoclast-cell rich sarcoma | 7 |
| Osteofibrous Dysplasia | 4 |
| PEComa | 8 |
| PHAT | 1 |
| Phosphaturic mesenchymal tumour (PMT) | 6 |
| Pleomorphic Sarcoma | 39 |
| PVNS/GCTTS | 1 |
| Rhabdomyosarcoma | 17 |
| Solitary fibrous tumour | 1 |
| Spindle Cell Sarcoma | 23 |
