## Supplementary Table 2 for "Osteosarcoma: novel prognostic biomarkers using circulating and cell-free tumour DNA"

| Tissue type | Number of cases | Disease status | Dataset |
| --- | --- | --- | --- |
| Adrenal gland | 80 | Cancer | TCGA-ACC (n=80) |
| Bile duct | 36 | Cancer | TCGA-CHOL (n=36) |
| Bladder | 412 | Cancer | TCGA-BLCA (n=412) |
| Blood | 62 | Cancer | GES20945 (n=6), GSE37965 (n=15), GSE39141 (n=29), GSE40005 (n=12) |
| Blood | 1067 | Healthy | GSE32148 (n=20), GSE40005 (n=12), GSE40279 (n=656), GSE41114 (n=2), GSE41169 (n=33), GSE42861 (n=335), GSE42865 (n=9) |
| Blood | 446 | Other disease | GSE32148 (n=28), GSE41169 (n=62), GSE42861 (n=353), GSE42865 (n=3) |
| Bone | 3 | Cancer | GSE38240 (n=3) |
| Bone marrow | 196 | Cancer | TCGA-LAML (n=140), GSE40870 (n=48), GSE42119 (n=8) |
| Bone marrow | 3 | Normal | GES20945 (n=3) |
| Brain | 870 | Cancer | TCGA-GBM (n=142), TCGA-LGG (n=516), GSE32283 (n=62), GSE36278 (n=136), GSE42882 (n=14) |
| Brain | 154 | Healthy | GSE41826 (n=145), GSE42882 (n=9) |
| Breast | 797 | Cancer | TCGA-BRCA (n=789), GSE29290 (n=8) |
| Breast | 8 | Normal | GSE29290 (n=8) |
| Cervix uteri/ovary | 818 | Cancer | TCGA-CESC (n=307), TCGA-OV (n=10), TCGA-UCEC (n=444), TCGA-UCS (n=57) |
| Colon/rectum | 428 | Cancer | TCGA-COAD (n=284), TCGA-READ (n=98), GSE29290 (n=6), GSE32283 (n=18)*, GSE42752 (n=22) |
| Esophagus | 185 | Cancer | TCGA-ESCA (n=185) |
| Eye | 80 | Cancer | TCGA-UVM (n=80) |
| Fetal/embryo | 9 | Normal | GSE31848 (n=5), GSE32362 (n=4) |
| Head/neck | 528 | Cancer | TCGA-HNSC (n=528) |
| Kidney/adrenal gland | 839 | Cancer | TCGA-KICH (n=66), TCGA-KIRC (n=319), TCGA-KIRP (n=275), TCGA-PCPG (n=179) |
| Liver | 445 | Cancer | TCGA-LIHC (n=377), GSE32079 (n=68) |
| Lung | 989 | Cancer | TCGA-LUAD (n=461), TCGA-LUSC (n=372), TCGA-MESO (n=87), GSE36216 (n=69) |
| Lymphoma | 48 | Cancer | TCGA-DLBC (n=48) |
| Pancreas | 184 | Cancer | TCGA-PAAD (n=184) |
| Prostate | 507 | Cancer | TCGA-PRAD (n=498), GSE38240 (n=9) |
| Sarcoma | 731 | Cancer | TCGA-SARC (n=261), TCGA-SKCM (n=470) |
| Stomach | 395 | Cancer | TCGA-STAD (n=395) |
| Thymus | 124 | Cancer | TCGA-THYM (n=124) |
| Thyroid gland | 507 | Cancer | TCGA-THCA (n=507) |
| Tongue/Tonsil | 90 | Cancer | GSE38266 (n=42), GSE38271 (n=6), GSE41114 (n=42) |
| Tongue/Tonsil | 2 | Healthy | GSE41114 (n=2) |

Supplementary Table 2 – Marmal-Aid and TCGA datasets (450K array data)

* 6 of these failed ChAMP normalisation: “"GSM799858" "GSM799859", "GSM799860"

"GSM799861", "GSM799864" "GSM799865"
